# Design, Development, and Usability Evaluation of a Voice App Experience for Heart Failure Management

**DOI:** 10.1101/2022.03.30.22273183

**Authors:** Antonia Barbaric, Cosmin Munteau, Heather J. Ross, Kuo-Cheng Tong, Joseph A. Cafazzo

**Affiliations:** Institute of Biomedical Engineering, University of Toronto, Toronto, ON, Canada; Centre for Global eHealth Innovation, Techna Institute, University Health Network, Toronto, ON, Canada; Department of Computer Science, University of Toronto, Toronto, ON, Canada; Ted Rogers Centre for Heart Research, Peter Munk Cardiac Centre, University Health Network, Toronto, ON, Canada; Department of Medicine, University of Toronto, Toronto, ON, Canada; Institute of Health Policy, Management and Evaluation, Dalla Lana School of Public Health, University of Toronto, Toronto, ON, Canada

## Abstract

The use of digital therapeutics (DTx) in the prevention and management of medical conditions has increased through the years with an estimated 44 million people using one as part of their treatment plan in 2021, nearly double the amount from last year. DTx are commonly accessed through smartphone apps, but offering these treatments through an alternative input can improve the accessibility of these interventions. Voice apps are an emerging technology in the digital health field, and may be an appropriate alternative platform for some patients. This research aimed to identify the acceptability and feasibility of offering a voice app as an alternative input for a chronic disease self-management program. The objective of this project was to design, develop, and evaluate a voice app of an already existing smartphone-based heart failure self-management program, *Medly*, to be used as a case study. A voice app version of *Medly* was designed and developed through a user-centered design process. We conducted a usability study and semi-structured interviews with representative end users (n=8) at the Peter Munk Cardiac Clinic in Toronto General Hospital to better understand the user experience. A *Medly* voice app prototype was built using a software development kit in tandem with a cloud computing platform. Three out of the eight participants were successful in completing the usability session, while the rest of the participants were not due to various errors. Almost all (7 out of the 8) participants were satisfied with the voice app and felt confident using it. Half of the participants were unsure about using the voice app in the future, though. With these findings, design changes were made to better improve the user experience. With rapid advancements in voice user interfaces, we believe this technology will play an integral role when providing access to DTx for chronic disease management.

## Introduction

### Background

The prevalence of heart failure (HF) continues to be on the rise as more people are surviving cardiovascular disease (1). Cardiovascular diseases can cause the heart muscle to become damaged and weak, leading to the development of HF. HF occurs when the pumping action of the heart muscle is not strong enough to meet the needs of the body, or when the heart muscle does not relax properly to accommodate blood flow back into the heart. When this occurs, fluid can build up in the lungs and other parts of the body, such as the ankles, creating congestion in the lungs and results in a lack of oxygen being delivered to the rest of the body (2). As of 2017, it is estimated that 64.3 million people are living with HF globally (3) and many countries are reporting a steady increase in this condition’s prevalence.

HF not only creates a burden on healthcare resources and expenditures (1), but also on the patient’s well-being if not cared for properly. HF limits a patient’s capacity to live well, either through physical, psychological, or social means (4). Patient self-management plays an integral role in the treatment of HF, with studies reporting improved health outcomes, decreased clinic visits, and decreased health costs (5). Mobile health, also referred to as mHealth, is a type of digital health technology that involves the use of mobile devices (smartphone, patient monitoring device, wireless devices, etc.) for medical and public health practice (6), and enables the integration of self-care support into a patient’s daily routine (7). Smartphone applications (apps) remain to be one of the most popular tools for helping patients who are diagnosed with chronic conditions manage their health at home (8).

*Medly* is an evidence-based, HF self-management program that has been developed by the University Health Network (UHN) and is implemented as part of the standard of care at UHN’s Peter Munk Cardiac Centre (PMCC) (9). This program is available to patients as a prescribed digital therapeutic, cleared by Health Canada in 2020. *Medly* consists of a smartphone app to enable patients to log clinically relevant physiological measurements and symptoms daily, which is then used in the *Medly* algorithm to generate an automated self-care message. The care team is able to review the patient’s data daily, and can view current trends and historic data. If the algorithm detects the patient is deteriorating, the care team receives alerts via email and on the dashboard, and is able to contact the patient to advise. Previous studies evaluating *Medly* have proven that this program can reduce health service utilization and improve clinical, quality of life, and patient self-care outcomes (9).

With these encouraging results, there is a desire to improve the accessibility of the *Medly* program. More specifically, there is evidence in the literature that suggests older adults, as well as those with cognitive and physical impairments have difficulties when using touch screen devices, such as the smartphone (10–13), and often require additional assistance from their caregivers, making it difficult to complete tasks independently (14–16). Similarly, a study performed by Ware et al. showcased that adherence rates for *Medly* over a 12 month period were highest in older age groups and progressively lower in younger age groups (17). This data suggests that younger adults may require a more convenient way of interacting with self-management apps that require consistent interaction. While smartphone design guidelines for these specific demographics exist (12), there is potential for other platforms, such as voice user interfaces (VUIs), to increase the uptake of this program and to create a better user experience.

Voice apps are an emerging technology in the healthcare field, ranging from integration with in-clinic registration processes to helping people manage their chronic illness and live independently in their homes. There is growing interest in VUIs such as Google Home and Amazon Alexa due to their simple set-up, ease of use, and low cost when compared to current interactive voice response technologies already on the market. Voice apps enable ubiquitous connectivity by allowing the user to access services using only their voice, making for a more convenient experience. Research relating to voice apps for chronic illness are still in the development and piloting phases and have limited efficacy in testing to support final outcomes and conclusions. As described by Sezgin et al., most voice apps currently being developed provide information and assistance services, which includes general educational content and guidance, as well as reminders and tracking (18). There is limited research showcasing voice apps as a tool to provide more personalized, user specific support, creating an opportunity to further investigate using this technology to provide healthcare services and support.

### Objectives

Guided by the findings from the literature review and coupled with a user-centered design process (UCD), we sought to design, develop, and evaluate a voice app for an existing smartphone-based HF self-management program, *Medly*, to determine if a voice app version adds benefit to *Medly*’s current model of interaction and care. The *Medly* voice app was evaluated in a usability study, and the findings from this evaluation helped inform the final design and development of the *Medly* voice app, as presented in this paper.

## Methods

### Overview

The *Medly* voice app was created in two main phases: 1) design and development, and 2) usability evaluation of the preliminary design. A prototype version of the *Medly* voice app was created and used in a usability study. Findings from the usability evaluation helped inform any redesign and redevelopment work needed to create a voice app that better met the needs of HF patients.

The *Medly* voice app was designed and developed for deployment on VUIs and was built using an external cloud computing service. The usability study was performed under the UHN REB 19-5051.2 and collected both quantitative and qualitative data regarding the user’s experience when interacting with the *Medly* voice app prototype. The findings from this usability study influenced any redesign and/or redevelopment work that occurred after the usability evaluation.

### Design Process

A UCD process was followed when the *Medly* voice app was created to ensure that end user needs were included in all phases of the design project. This framework is an iterative process that allows design teams to create technologies that users are able and willing to use, rather than expecting them to change their behaviors in order to accommodate the technology (19). The first step in this process is concept generation and ideation, which involves identifying end user needs to better understand the technology’s intended use. Following this investigation prototypes are developed, based on the design requirements created from the user needs assessment, and evaluated through usability testing. Multiple cycles of prototyping and user testing are performed to improve the ease of use and adoption of the technology.

For this research, a literature review was performed during the first phase of the UCD process to help guide the voice app design by better understanding the demographic that would use this technology and the environment in which it would be used. Keywords such as, “chronic illness”, “self-management”, and “Amazon Alexa’’ or “Google Home’’ were used in library database searches in order to find relevant research related to this project. A market scan was also performed in addition to the literature review and specifically searched for voice apps related to healthcare management. The ideation of the *Medly* voice app was also influenced by the *Medly* program requirements. Using the information gathered from the first phase, a prototype of the *Medly* voice app was developed and deployed on a VUI. A usability study with the voice app was then performed with representative end users in the Ted Rogers Centre of Excellence in Heart Function clinic at the PMCC in Toronto General Hospital. Based on the usability study results, the prototype was further developed and redesigned to better meet end user needs, and is expected to be evaluated in a future clinical study. Fig 1 illustrates the process that was followed for this project.

**Fig 1.**
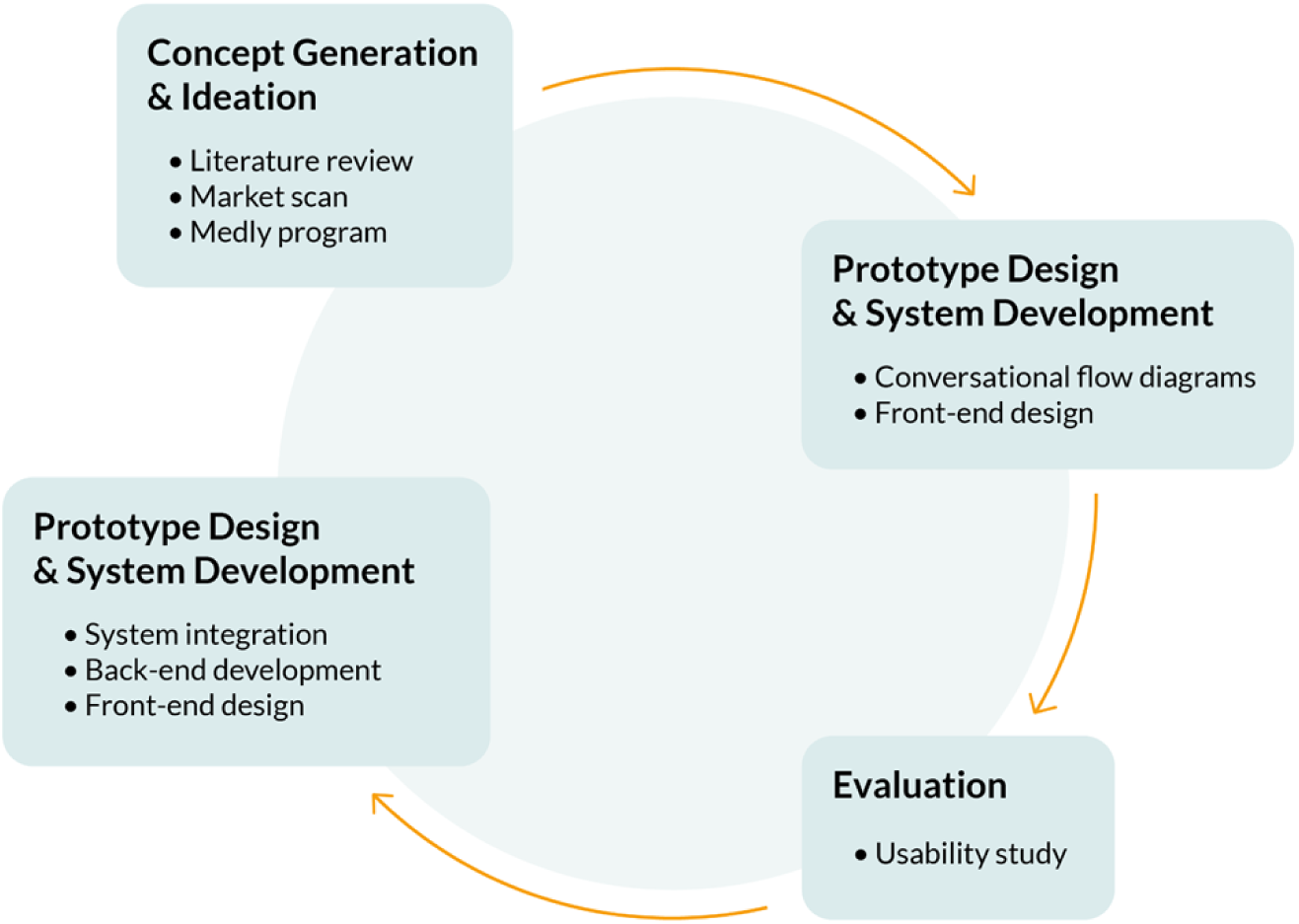
Design process was guided by UCD process; details describing what was done during each phase are included above.

### Design Specifications of Medly *Voice*

Design requirements were created prior to developing the *Medly* voice app to help guide the development process and define the voice app’s functionality. The main requirements implemented for the voice app are presented in Table S1 in Multimedia Appendix 1, which summarize the main objectives of the program. The voice app was designed to have the same functionality as the mobile app which includes: 1) asking the user to measure their weight, blood pressure, and heart rate; 2) saving and storing those values in the *Medly* clinical dashboard; 3) asking the user a series of yes/no questions relating to HF symptoms; 4) processing the data using the *Medly* algorithm; and 5) outputting a message to the user based on the algorithm result. These requirements helped create an app that was appropriate for voice interaction and are based on research as well as guidelines related to VUI design (20,21). Conversational flow diagrams were created by also using VUI design guidelines, and each scenario was tested on a VUI following its implementation (22,23).

### Development Specifications of Medly *Voice*

#### Medly Voice App Architecture (Fig 2)

The architecture of the *Medly* Voice program consists of various components required to receive, store, and send data. The *Medly* voice app serves as the primary point of contact for the user and was built using a software development kit. The voice input of physiological measures is captured by the *Medly* voice app and is processed through an external serverless computing platform into algorithm input data. This data is transmitted to the UHN *Medly* Voice application server. This server receives the input data from Amazon Web Services, pulls relevant patient parameter thresholds from the *Medly* Voice application server, and evaluates the combined data using the *Medly* algorithm to generate a patient assessment. The patient data and resulting assessment are sent to the UHN *Medly* Application server, where it is stored in a database and available to view on a dashboard by the *Medly* care team.

**Fig 2.**
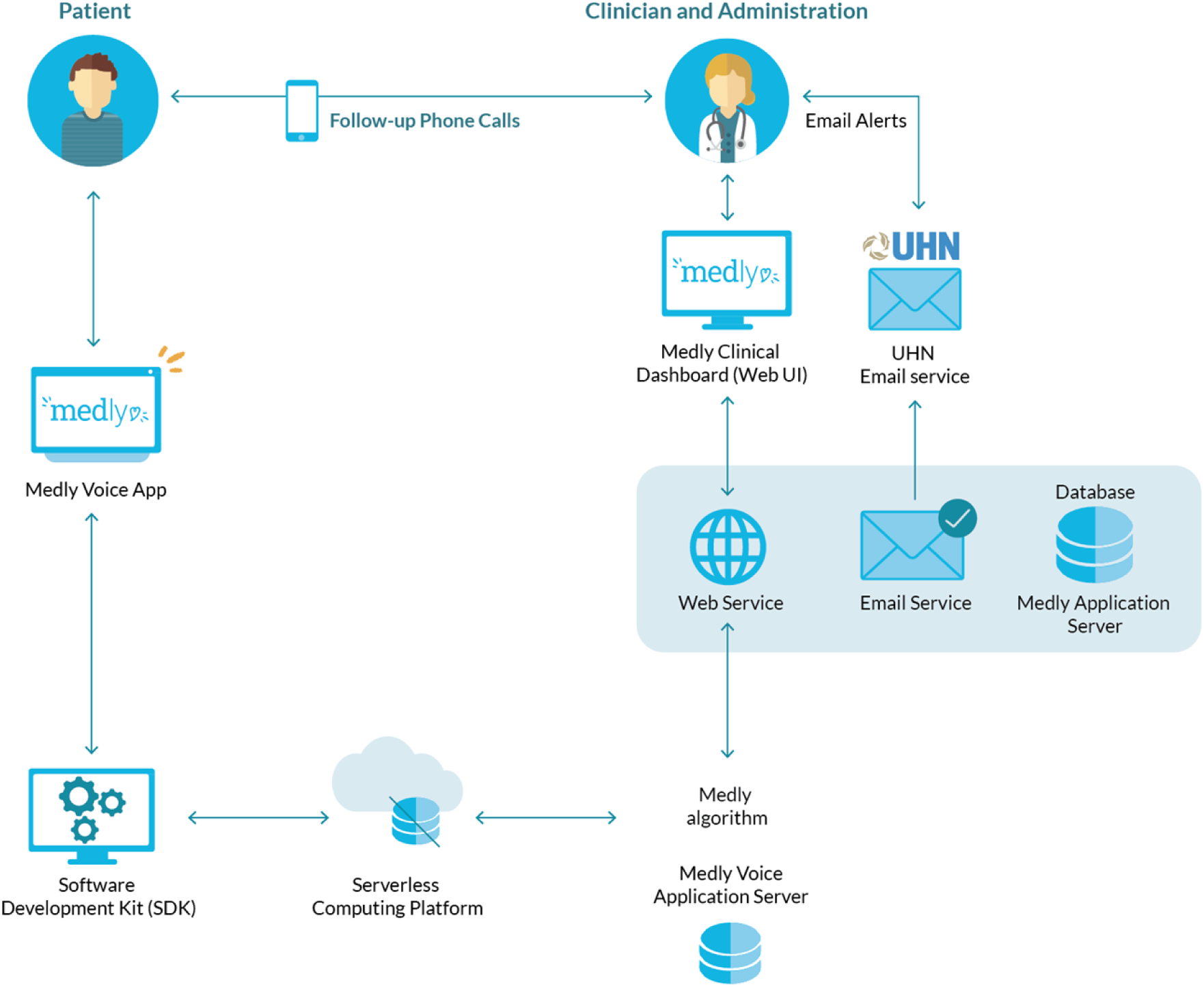
Software architecture diagram of the Medly voice app system. The participant interacts with the Medly voice app through the smart speaker, which is connected to an external cloud computing platform. This data is then sent to the Medly voice application.

#### Software Platform

Voice apps consist of both a VUI and application logic. When the user speaks to the VUI and invokes the voice app, a request is created and processed in the context of the voice app’s interaction model using tools such as machine learning, natural language understanding, and automatic speech recognition. Once the interaction model has processed the speech, a request is sent to the application logic, which provides a response back to the user. For the purposes of this project, a Hosted Alexa Skill built using the Amazon Alexa Skills Kit (24) was used to implement the *Medly* voice app’s logic, but other frameworks could have been used as well.

A voice interaction model consists of the following three inputs: 1) intents, 2) custom slot types, and 3) sample utterances. An intent represents an action that will fulfill a user’s request and uses slots to store key information extracted from the user’s request. Sample utterances are specific words and phrases that the user may say to invoke an intent. Developers are encouraged to include multiple sample utterances per intent to increase the likelihood of the interaction model entering into that intent when appropriate. Machine learning and natural language understanding are also used to help train the interaction model in identifying which intent to enter based on the phrase, even if it does not exactly match the sample utterances listed.

#### Verification and Validation

The verification and validation of the *Medly* voice app were ongoing processes that were completed at various steps in the design process. The verification process involved both the voice app code (25), as well as the *Medly* algorithm. The validation process consisted of evaluating the front-end of the voice app, which is what the user is expected to interact with.

Verification was done on the *Medly* voice app to ensure that it met the design requirements and performed as expected. The code implemented to help guide the user through the conversation was verified using a demonstration method. The demonstration method is one of the four fundamental approaches used in design verification, and can be appropriate for designs that consist of software development. Using the demonstration method, different scenarios were created with various inputs in order to ensure that the produced results were as expected. The *Medly* algorithm was verified using unit testing to ensure that the algorithm outputs the expected response. Various combinations of weight, blood pressure, heart rate, and symptom responses were designed using a test *Medly* patient account to determine if the correct self-care messages were being produced.

Different conversational flows were brainstormed and used to help validate the voice app. These conversations were then tested using an online voice app simulator, which responds with voice and text and also includes the JSON request to help fix errors. In addition to validating the *Medly* voice app by testing different conversational flows, the usability study helped determine the different conversations/phrases being used that were originally unaccounted for. These findings were then used to help inform the re-development work which included modifying the interaction model.

### Usability Study

#### Overview

The purpose of the usability study was to investigate the potential of voice communication devices as an alternative healthcare delivery platform to smartphone apps for providing patient self-care. Representative end-users were recruited and asked to explore the intuitiveness of the *Medly* voice app. Through the evaluation we sought to identify user preferences and expectations through the following: a usability session, the standardized System Usability Scale (SUS) questionnaire (Multimedia Appendix 3), and a qualitative semi-structured interview session. The findings from this study helped identify the acceptability and ease of use of the voice app and also informed the service design that would be most appropriate to implement for a voice app offering a self-management program.

#### Participant Recruitment

Eight participants were recruited for this study, and all of them use *Medly* as part of their standard of care. All eight participants recruited were adults living with chronic HF and could read, write, and understand English. *Medly* patients were asked to participate so that unique insights and themes could be identified since they already have experience with the program. Participants were recruited through the *Medly* nurse coordinator, who provided a brief overview of the research study and then asked if they were interested in participating before introducing them to the study coordinator. Most usability studies require 5-8 participants in order to receive response saturation (26,27) and as a result we chose to recruit eight participants to help identify the potential this platform may have as a healthcare delivery technology in the future, by identifying major themes emerging from the study.

#### Usability Study Design

Participants who volunteered attended a single, one hour session with the study coordinator after their scheduled appointment and were asked to complete four main tasks: 1) inputting measurements on *Medly* smartphone app, 2) inputting measurements on *Medly* voice app, 3) completing the SUS questionnaire, and 4) engaging in a semi-structured interview with the study coordinator. The first task asked the participant to interact with the *Medly* app on a phone provided by the study coordinator. While the participant was completing this task, the study coordinator recorded observational notes. Following this, the participants were then asked to verbally interact with the *Medly* voice app using a VUI, which was provided by the study coordinator. The user was also provided with an instruction card with steps to follow that included suggestive phrasing to help guide them through the conversation. During this time, the facilitator also recorded observational notes. Participants then completed a standardized, post-study SUS questionnaire regarding their experience with the voice app. Once participants completed the questionnaire, the study coordinator led a semi-structured interview with the patient which included questions pertaining to their perceptions of the usability of the *Medly* voice app, and specific issues or areas for improvement. The results of this research highlighted key findings regarding the acceptability of integrating voice communication platforms into patient-self-care tools, and informed decisions for future design iterations.

## Results

### Preliminary Design of Medly Voice App

A voice app prototype of *Medly* was created for the purpose of it being used during a usability study in order to identify problems in the design, uncover any opportunities for improvement, and to learn more about the users’ behaviors and preferences when interacting with the app. An instructions card was created and used to help guide the user through the *Medly* voice app interaction (Figure S1, Multimedia Appendix 1). A preliminary design of the card was used by participants in the study so that feedback could be collected about the design to make changes, if required.

A high level overview of the conversation was first mapped out and created (Fig 3). Using this, the full conversation was designed and is described in more detail in Figure S2 in Multimedia Appendix 1. The first half of the voice app interaction consisted of the user measuring and recording their weight, blood pressure, and heart rate with non-lyrical music playing in the background in between prompts. Playing music in the background not only created a more pleasant user experience, but was also a strategy used to ensure the app did not time out (28). After the user inputted their heart rate, the voice app reiterated the measurements it captured to the user and gave them an opportunity to correct any wrong measurements. Once the user recorded their measurements, the app began to ask them a series of yes/no questions of HF-related symptoms. Once complete, the app outputted a message and exited. The *Medly* algorithm was not implemented for the prototype because it was not needed to accomplish the purpose of the usability study.

**Fig 3.**
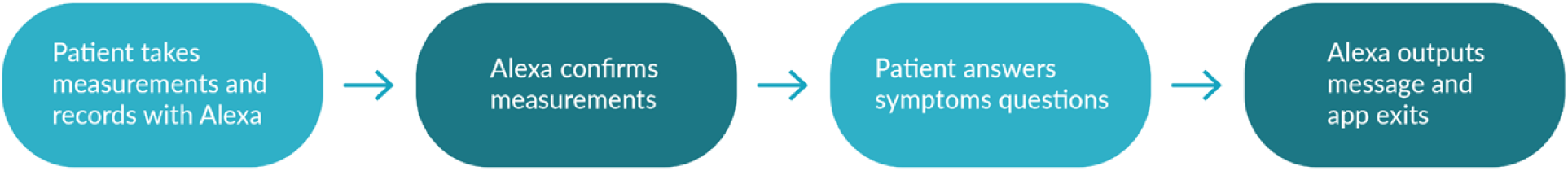
A high level description of the conversational flow for the Medly voice app. ‘Milestones’ are outlined and signifies an important event.

An interaction model was then created to implement the conversational flow described above and consisted of intents, training phrases, and slot types. An intent was created for each piece of required information: weight, blood pressure, heart rate, and symptom questions. Within each intent, training phrases were added and consisted of predictable utterances the user may say to record their responses. Within each training phrase, the most important pieces of data were identified and labeled as a slot type (weight, blood pressure, heart rate categorized as ‘numbers’, with symptom responses labeled as ‘yes’ or ‘no’). The slot types were used to help extract the measurements needed to send to the *Medly* voice server. Measurements were extracted from the interaction model and the application logic was used to help direct the flow of conversation. Once the required measurements were captured, a POST request was made to the *Medly* server, and a self-care message sent back to the voice app to relay to the user.

### Usability Study Findings

A usability study was performed with eight HF participants from UHN’s Ted Rogers Center of Excellence for Heart Failure clinic. Three out of the eight participants were able to successfully complete the usability session, while the other participants were unsuccessful due to the voice app exiting because of various errors that occurred, mainly with the device unsuccessfully understanding numerical values the user would say. Despite these results, all participants filled out the SUS questionnaire to describe their experience with the voice app. The responses from this questionnaire resulted in an average SUS score of 92 (out of 100), ranking the voice app in the 98^th^ percentile based on previous studies. Most participants were satisfied with the design and development of the voice app. Participant responses for statements relating to the positive and negative attributes of the voice app are shown in Figs 4a and 4b, respectively.

**Fig 4.**
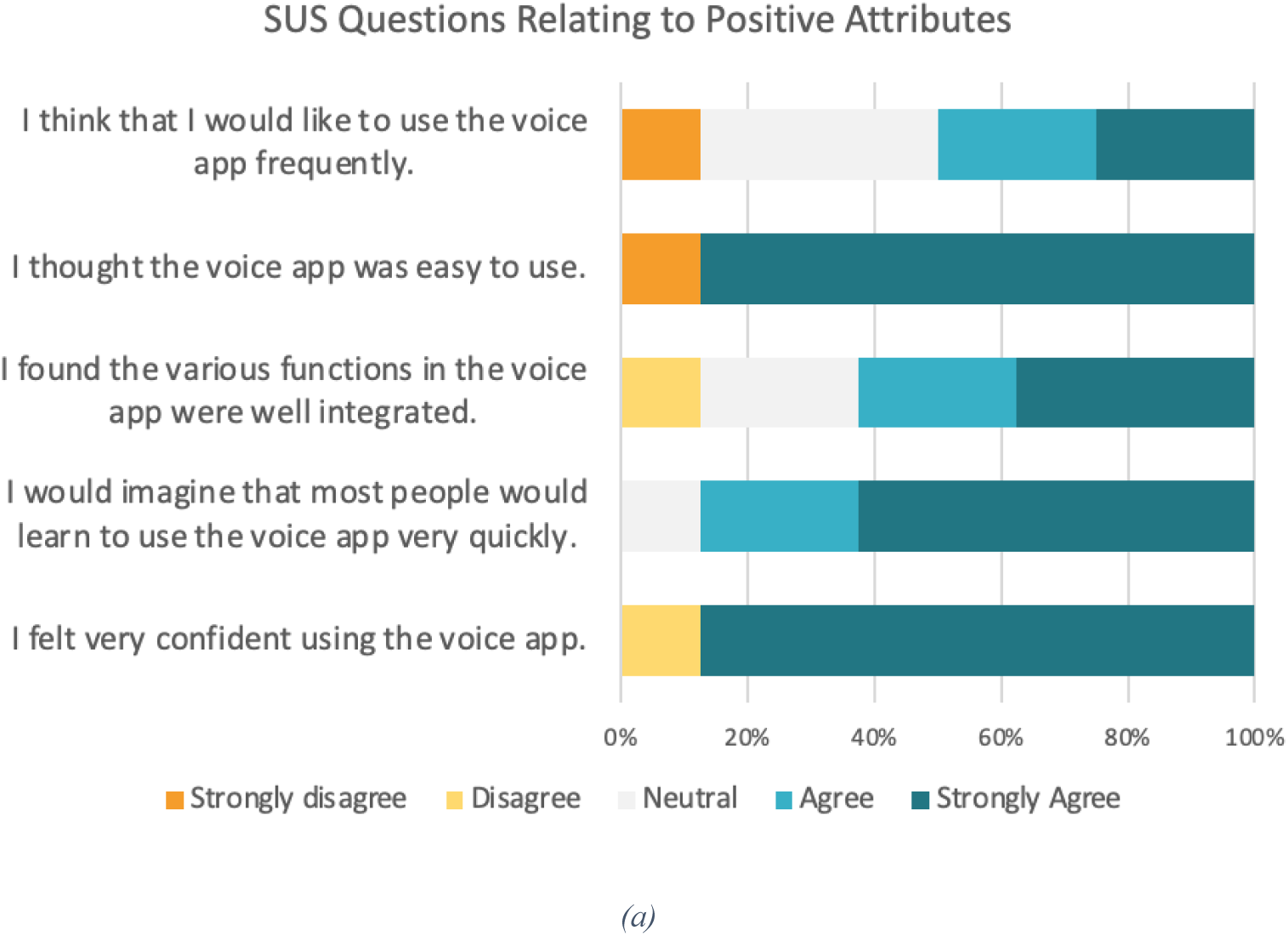
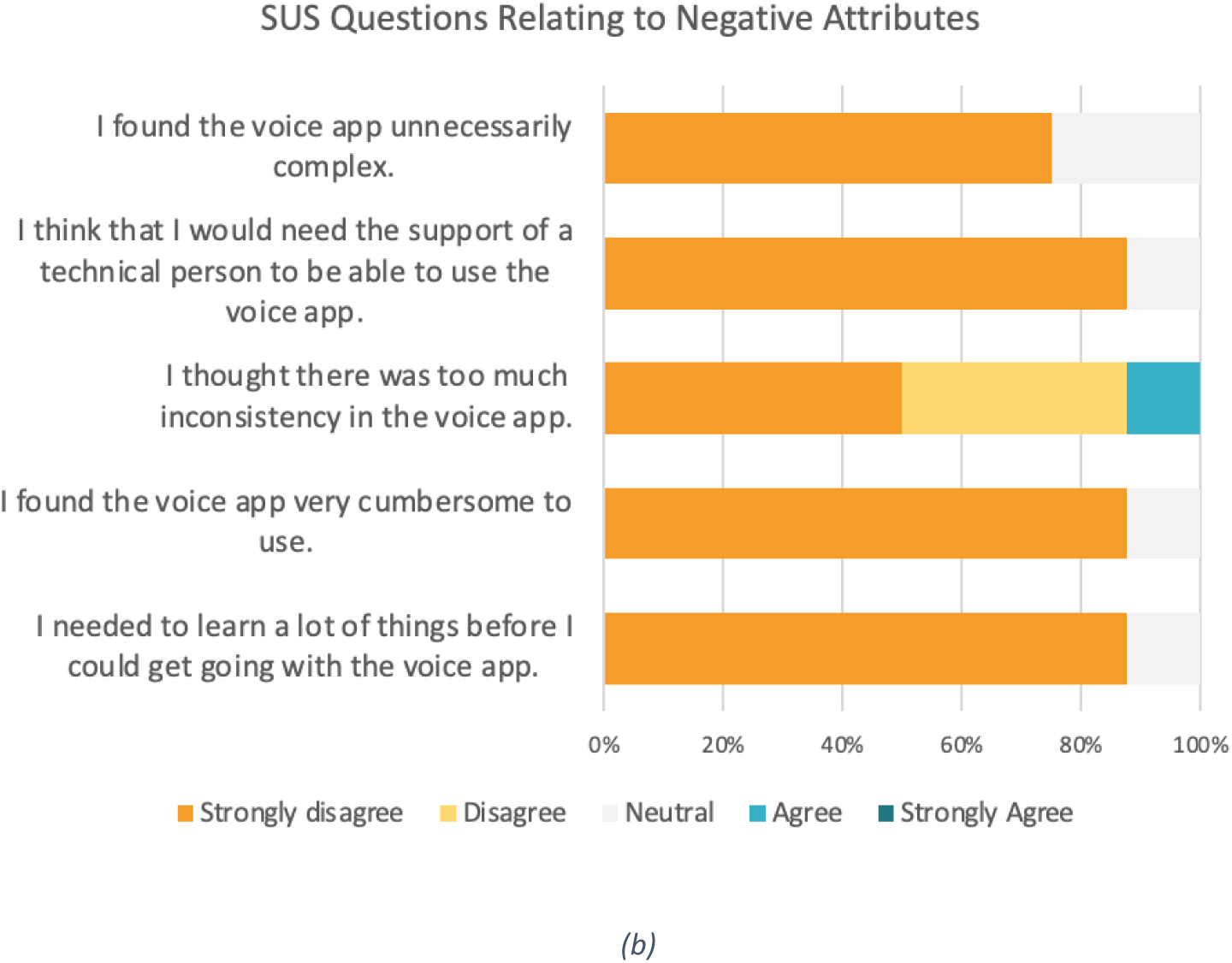
(a) Data showcasing participant results for the questions in the SUS relating to positive attributes of the Medly voice app system (b) Data showcasing participant results for the questions in the SUS relating to the negative attributes of the Medly voice app system.

Looking at the positive attribute statements, almost all participants (7 out of the 8) felt confident using the app and thought it was easy to use. This is further supported with the responses to the negative attributes statements, with the most popular response being ‘strongly disagree’. When asked about whether they would use the voice app frequently however, there was a greater divide in opinion, with the most popular response being neutral.

Overall, these findings show promise when determining user acceptability due to the overall positive responses from the SUS questionnaire. These results also indicate that some minor changes should occur in order to improve the patient expereince. Participants who were unable to complete the session had difficulties recording their blood pressure readings, either due to the way they said the readings (saying each digit separately for systolic and diastolic), or if they took a small pause between values, in which case the voice app incorrectly recorded the values. In these cases the users were not able to correct the measurements. As a result, the blood pressure part of the interaction model was further developed to mitigate the risk of incorrectly and/or unsuccessfully capturing the user’s data.

In addition to the SUS questionnaire results, observations were derived from the semi-structured interviews that occurred with each user after their usability session. For example, an obvious change in physical behaviour was observed with participants when interacting with the voice app, with most users showcasing nervous and tense feelings during the session. Furthermore, some voice app features helped the user navigate through their session making for a more pleasant experience, such as: having the music play in the background so they didn’t feel rushed through the interaction, having the voice app reiterate measurements back to them to ensure the device correctly recorded the values, as well as having the instructions card available with suggested phrases to use. General frustrations were expressed by participants when using a smartphone, and concerns about privacy were brought up by two (out of eight) participants. These findings are described in more detail in Table S1 in Multimedia Appendix 2.

Prior to the second part of the development phase, a list of potential risks were identified that relate to the user interacting with the voice app. The severity of each risk was assessed based on its likelihood and consequence. Mitigation strategies for each risk were developed and implemented in the voice app design during the redevelopment work. This data can be seen in Table S2, Multimedia Appendix 2.

### Redevelopment of *Medly* Voice App

The prototype that was built for the usability study was further developed to meet all of the *Medly* voice app design requirements. The new features added were influenced by the findings from the usability study and also involved changing VUIs to incorporate screen and touch screen capabilities.

#### Screen Design

Having a screen display on the device meant that users were able to visually see instructions and measurements, and had the option to respond using the touch screen for yes/no questions. Each screen was designed to only include the necessary information the user needed to avoid any confusion. A consistent theme, similar to the *Medly* smartphone app, was used for each screen display by using the same color scheme, layout, font size and style. The written cues on the screen aligned with the verbal prompts to avoid user confusion. Design guidelines were used for font size and layout to ensure the prompts are presented at an appropriate reading size (29). The screen design at various points of the conversation is showcased in Fig 5.

**Fig 5.**
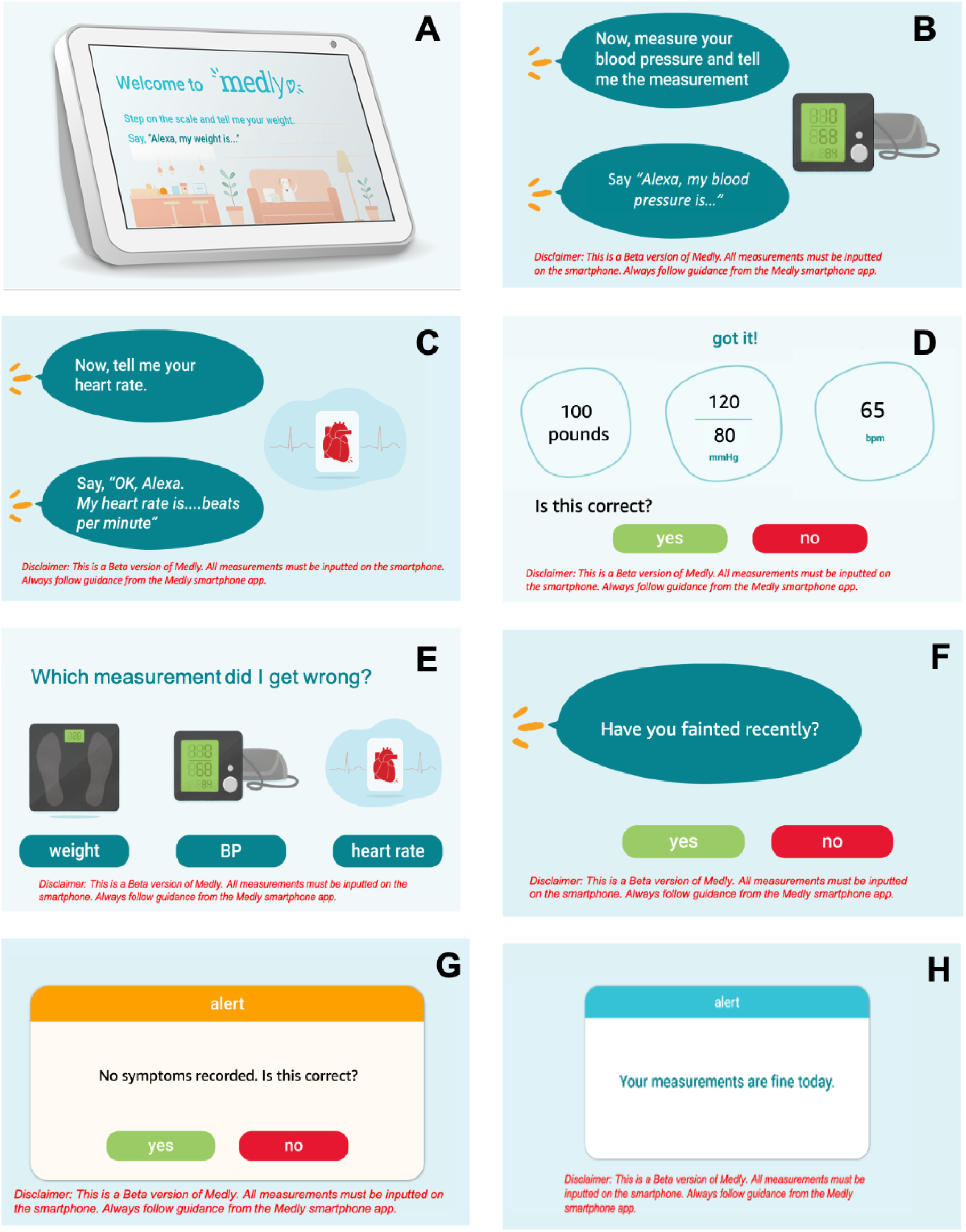
Screen designs showcased through images A-D, representing different events that occur in the conversation. A disclaimer is said verbally at the start of skill interaction, and in between a written disclaimer is provided on the screen. (A) welcome screen that users see when they invoke the skill, (B) and(C) instructions to measure blood pressure and heart rate, respectively, (D) confirmation slide of the user’s weight, blood pressure, and heart rate, (E) what the user sees when they are correcting a measurement, (F) an example of how the symptom questions appear on the screen, (G) the confirmation message for the symptom responses, (H) an example of the output message provided by the Medly algorithm.

Incorporating a screen display for the *Medly* voice app would also allow users to see their previous measurements and data trends, similar to what they’re used to seeing when using the smartphone app. While the implementation of seeing data history was not in scope for this project, it is a feature that is common in most mHealth apps since it helps users understand their health status better.

#### Conversational Design

The conversational script used in the preliminary design stayed the same during the re-development work. Findings from the usability study helped inform which parts of the conversation were most prone to error and needed to be changed. More training phrases and symptom confirmations were new additions to the conversation and the summary of the interaction is shown in Fig 6. A more detailed description of the new additions to the conversation can be seen in Figure S3 in Multimedia Appendix 1. With these new changes, the app only confirms the symptoms the user answered yes to for a more efficient process by giving them an opportunity to correct any wrong responses recorded. Once all the responses have been corrected, the data was sent to the *Medly* voice server. The voice server retrieved patient thresholds, sent the daily data to the *Medly* research server and used the *Medly* algorithm to send an output message back to the voice app, informing the patient on the status of their health.

**Fig 6.**
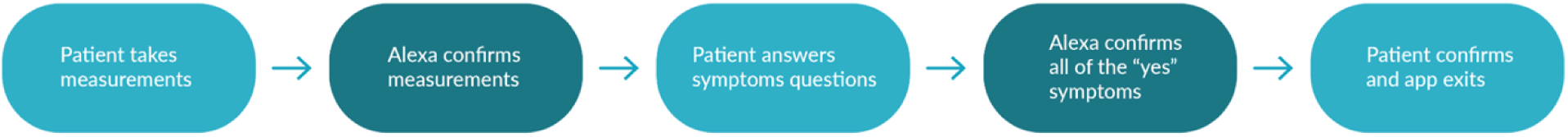
A high level description of the conversational flow for the final version of the Medly voice app. A new milestone has been added to describe Medly voice app also confirming the symptom responses answered ‘yes’.

#### Back End Development

The software architecture and *Medly* algorithm were implemented during the redevelopment phase of the project. Each output message generated to the user was a personalized response based on their measurements and pre-existing thresholds. In order to receive data from the voice app and pull relevant patient thresholds, account linking was required to be set-up to connect the patient’s identity from the two different services. Personalized *Medly* accounts were created for each study participant and connected to their respective Amazon account. The *Medly* algorithm was built and deployed on the *Medly* voice server and pulls data from *Medly* application server as well as the data sent from the VUI to generate an output message.

## Discussion

### Principal Findings

This manuscript describes the UCD approach that was used to develop a voice app experience for HF management. Accessing chronic disease management programs through the use of voice apps has the potential to increase these programs’ uptake by making them more accessible. The results from this usability study show promise in the possibility of offering these types of programs as a voice app, and the development process proves the feasibility of deploying these programs on VUIs.

VUIs are designed to be integrated within a household by having the ability to connect to multiple devices throughout the home (“smart devices”), and also provide the ability to connect to online services by just using voice. Their near ubiquitous nature makes it easy and convenient to perform tasks through simple conversation and offers the flexibility to do other tasks simultaneously. Patients with chronic illness experience a constant need to record and transmit data to better track their health and VUIs have the potential to help them accomplish this seamlessly, not only because it uses speech to communicate but also because it can help guide the user through their tasks more explicitly than the smartphone.

Because this platform requires less technical knowledge than the smartphone (mHealth apps), those who struggle to use technology have the opportunity to participate in the program. Based on the usability study observations, most participants felt that it was easy to interact with the voice app since it only required a conversation and no technical background. The platform’s multilingual capability offers another opportunity, which may make some more willing to log their measurements. VUIs also allow users to initiate a call to anyone on their list through voice command, which can be useful in cases where they need assistance but cannot reach their phone to call for help. This scenario often occurs with older demographics who are also the most common group to be diagnosed with chronic illnesses, and as a result are prescribed digital therapeutics.

When inputting measurements on a mHealth app, it is typical for the user to see their data on the screen prior to submitting. With a VUI that solely relies on verbal communication, visual feedback is no longer a possibility. Study participants commented on the importance of knowing which measurements were recorded on the voice app, and valued the ability to see historical data and trends on the smartphone app. With these findings, we incorporated verbal feedback as a necessary component in the conversational design to help with user confidence. Adding this step in the conversation comes at the cost of prolonging the time it takes to complete the interaction, but is not necessarily a drawback due to the lower focus required when compared to using a smartphone. Advancements in VUI technology include the ability for the user to interrupt the device when speaking, and an integrated screen display which can allow the users to view results (as they are currently accustomed to), as well.

Although users only need to use speech when interacting with VUIs, the conversation style may vary significantly when compared with human-to-human interaction. During the study, users who had never used these types of devices before felt the need to raise their voice, in hopes that it heard and understood them better. Expelling this energy repeatedly may make the user feel tired, as one participant noted, especially since the demographic who uses these digital therapeutics are older adults. Without knowing how the voice app will respond, a difference in body language was also observed, namely users would tense up, felt the need to sit up straighter, and focused more when interacting with the device. This behaviour is in stark contrast to how users interacted with the smartphone, since the physical body language requires a much more relaxed behaviour. Despite these differences, it is expected that user body language will shift towards a more relaxed behaviour as they get more comfortable using voice apps.

It is also important to consider the potential barriers to entry that exist as a result of using this technology. For example, most VUI devices require the user to have an account to interact with the device, creating an accessibility issue. Another barrier to entry is the requirement to have a constant, reliable internet connection to use the device. This is in contrast to the *Medly* smartphone app which can be completed offline and is mobile. Not only can this create an equity issue for those who do not have internet, but can also be problematic when internet issues occur and power outages.

As advancements in VUIs progress, we believe they will play an integral role in providing access to chronic disease management programs, by: 1) helping more patients complete their tasks independently, 2) offering a more convenient experience to record relevant data, and 3) allowing those with limited technical and English skills access to these programs.

## Limitations

While the results from this research project show promise for future use, it is also important to acknowledge the limitations associated with this work.

The main limitation for usability studies is the short interaction time. While the observations and feedback from participants was helpful in identifying the potential voice apps may have for chronic disease management, they only interacted with the technology for a short period of time. Future work should include a study that requires the users to interact with the device for a longer period of time to give them more experience with the voice app to reflect more grounded feedback.

A sample size of eight participants was chosen since the usability study was only focused on gathering user insights, and not statistically relevant data. According to the literature, the total participants needed for a usability study is dependent on when saturation in results is achieved. In most cases this occurs when the sample size is typically between 5-8 participants (30). Therefore, it is recommended that multiple usability tests occur with fewer users (between 5-8 participants) and that changes be made between usability tests to mitigate challenges observed in previous sessions. The research showcased in this paper underwent 11/2 cycles of the UCD process and requires further testing to identify whether the challenges uncovered in the usability study are resolved with the latest iteration of design and development work. Due to the small sample size, the usability study is also not statistically powered. In this case, the evaluation of the results is mostly limited to qualitative data analysis. While a questionnaire was completed by each user after the session, these results are only used to support the conclusions drawn from the qualitative data.

Lastly, participants may have had social desirability bias when answering interview questions and filling out the questionnaire, especially because they knew that the study coordinator also had involvement in the design and development of the voice app (31). When things go wrong, participants most often feel as though it is their fault for not understanding how to use the technology, and as a result will not bring up comments about what they disliked about the experience since they think that it is their fault.

## Conclusion

This project involved the design, development, and usability evaluation of a voice app for HF management. A UCD process was followed to systematically create a voice app that would meet user needs and be easy to use. The usability study performed at the PMCC at the University Health Network provided insightful user feedback about the voice app design, with the overall response being positive with high user satisfaction. The findings from this usability study impacted the re-development of the voice app, which will be used in a future clinical evaluation. The findings from this research show promise in using VUIs to help with chronic disease management.

## Data Availability

The work that was produced and collected in this study is stored in University Health Network (UHN) servers. Our agreement with the UHN Research Ethics Board describes very specific circumstances that does not allow us to make the data publicly available and prohibits us from sharing the data on a public platform.

## Acknowledgments

The authors wish to thank Jacqueline Simpson for designing the *Medly* voice app screensavers and figures presented in this paper. The authors are also grateful to all of the patients who gave their time and participated in this study.

## Abbreviations

HF: heart failure
PMCC: Peter Munk Cardiac Center
SUS: System Usability Scale
UCD: user-centered design process
UHN: University Health Network
VUI: voice user interface

## Supporting Information

Multimedia Appendix 1 Table. S1

Multimedia Appendix 1 Fig. S1

Multimedia Appendix 1 Fig. S2

Multimedia Appendix 2 Table. S1

Multimedia Appendix 2 Table. S2

Multimedia Appendix 3 Fig. S1

